# Spectrum of neurological manifestations and systematic evaluation of cerebrospinal fluid for SARS-CoV2 in patients admitted to hospital during the COVID-19 epidemic in South Africa

**DOI:** 10.1101/2021.05.14.21254691

**Authors:** Angharad G Davis, Marise Bremer, Georgia Schäfer, Luke Dixon, Fatima Abrahams, Rene T Goliath, Mpumi Maxebengula, Alize Proust, Anesh Chavda, John Black, Robert J Wilkinson, And the HIATUS Study Consortium

**Affiliations:** Wellcome Centre for Infectious Diseases Research in Africa, University of Cape Town, Observatory 7925, Republic of South Africa; The Francis Crick Institute, Midland Road, London, NW1 1AT, United Kingdom; Faculty of Life Sciences, University College London, WC1E 6BT, United Kingdom; International Centre for Genetic Engineering and Biotechnology (ICGEB), Observatory, Cape Town, 7925, Republic of South Africa; Imperial College NHS Healthcare Trust, Charing Cross Hospital, London, W6 8RF, United Kingdom; Department of Infectious Diseases, Imperial College London, London, W12 0NN, United Kingdom; Department of Radiology, West Middlesex University Hospital and Chelsea and Westminster Hospital, London, United Kingdom; Department of Medicine, Walter Sisulu University, Mthatha 5117, Republic of South Africa

## Abstract

Neurological manifestations of COVID-19 are increasingly described in the literature. There is uncertainty whether these occur due to direct neuroinvasion of the virus, para-infectious immunopathology, as result of systemic complications of disease such as hypercoagulability or due to a combination of these mechanisms. Here we describe clinical and radiological manifestations in a sequential cohort of patients presenting to a district hospital in South Africa with neurological symptoms with and without confirmed COVID-19 during the first peak of the epidemic. In these patients, where symptoms suggestive of meningitis and encephalitis were most common, thorough assessment of presence in CSF via PCR for SARS-CoV2 did not explain neurological presentations, notwithstanding very high rates of COVID-19 admissions. Although an understanding of potential neurotropic mechanisms remains an important area of research, these results provide rationale for greater focus towards the understanding of para-immune pathogenic processes and the contribution of systemic coagulopathy and their interaction with pre-existing risk factors in order to better manage neurological disease in the context of COVID-19. These results also inform the clinician that consideration of an alternative diagnosis and treatment for neurological presentations in this context is crucial, even in the patient with a confirmed diagnosis COVID-19.

## Introduction

Although COVID-19, the disease caused by SARS-CoV2, is primarily a disease of the respiratory tract, coronaviruses (CoV) are known to be neurotropic with some strains leading to meningitis, encephalitis and cerebral vasculitis [20]. In the first case series of SARS-CoV2 conducted in Wuhan, China, it was found that in 62 patients, 21 (34%) reported headache [1]. In a subsequent retrospective case series to investigate neurological presentations in 214 patients with COVID-19, 36% of patients were found to have neurological signs and symptoms, which were more frequent in severe compared to non-severe COVID-19 (45% vs 30.2%). Since then, a wealth of literature suggests that SARS-CoV2 has the potential to present with neurological manifestations, with or without pulmonary involvement. Reports include cases of meningitis/encephalitis, with [2, 3] and without [4] detection of SARS-CoV2 within the cerebrospinal fluid (CSF), stroke [5], transverse myelitis [6-8], acute disseminated encephalomyelitis [9] and inflammatory polyradiculopathies such as Guillain-Barre Syndrome [10-13]. Whether the mechanism by which these phenomena occur are due to, either individually or in combination, direct neurotropic invasion, para- or post-infectious inflammation, a systemic coagulopathic processes or indeed due to separate and co-incidental pathogenic processes is not well understood.

The distinction between direct CNS invasion and other potential mechanisms is however important to make. Clinicians who assess patients presenting with neurological symptoms in the context of a COVID-19 epidemic must know when and how to investigate for neurological complications of SARS-CoV2 infection. Moreover, should SARS-CoV2 commonly lead to neurological symptoms via direct CNS invasion, particularly in the absence of pulmonary symptoms, then patients should be screened routinely for neurological involvement, and physicians must consider this unique potential of the virus. If direct SARS-CoV2 invasion to the CNS does not explain neurological presentations in this context, then greater focus must be towards understanding the nature and contribution of para- and post-infectious inflammatory phenomenon and coagulopathy in order to develop effective therapeutics to treat these often severe complications of disease. This is particularly important in resource limited settings where testing for SARS-CoV2 is not always available, and as yet there are no bespoke methods to test for SARS-CoV2 in cerebrospinal fluid (CSF). In these settings the precise incidence of SARS-CoV2 infection may not be known, or be underestimated: in the Eastern Cape of South Africa for instance where this study took place, seroprevalence following the first wave of the epidemic has been found to be as high as 62.5% in adults under the age of 65 years which is 8-fold higher than the official case count [14], and has led to death in an estimated 1 in 300 people in the region (SAMRC data) [15]. Such high rates of infection and subsequent mortality not only calls for better resource to manage the disease in these settings, but should also alert the clinician to the increased possibility of encountering less typical presentations of SARS-CoV2 infection.

To date no published studies have systematically investigated for the presence of SARS-CoV2 within the CNS in patients presenting with neurological symptoms with and without pulmonary manifestations of COVID-19 in a context where COVID-19 infection is the most frequent reason for hospital admission. In a prospective cohort study in the Eastern Cape of South Africa, we described clinical and radiological features, and assessed for the presence of SARS-CoV2 within the CNS in those presenting to hospital with neurological symptoms during the first peak of the COVID-19 epidemic.

## Methods

### Patient recruitment

We undertook a prospective cohort study at Livingstone Hospital, Eastern Cape. The study was approved by the Faculty of Health Sciences Human Research Ethical Committee of the University of Cape Town (HREC 207/2020) and by the ethical review board at Livingstone Hospital. We sequentially enrolled adults (>18 years) presenting with neurological symptoms who at the discretion of the treating physician required inpatient investigation by lumbar puncture and cerebrospinal analysis between 12^th^ July and 20^th^ October 2020. During this time Livingstone Hospital served as a COVID-19 referral centre in the Eastern Cape with an average admission rate of 40 confirmed COVID-19 cases per day. Written informed consent was taken from the patients where possible in those with capacity to consent. In those with decreased consciousness, patient relatives were approached for proxy consent. In those where no relative was contactable, permission was sought on an individual case basis by the Faculty of Health Sciences Human Research Ethical Committee of the University of Cape Town.

### Clinical and radiological data collection

Clinical data was collected on symptoms and signs at presentation as well as relevant past medical history at two timepoints (baseline, and again between 3 and 7 days of enrolment). Computerised Tomography (CT) head images performed as part of standard of care included as routine sagittal and axial views. Two dimensional images were retrieved and independently viewed using a picture archive and communication system (PACS) by a blinded neuroradiologist using a standardised case report form. No specific study procedures took place, however at the time of diagnostic lumbar puncture a total of 6 ml of additional CSF was retrieved for study specific analysis and biobanking. Similarly, venipuncture was not performed as a study procedure, however 24 ml of additional whole blood samples were collected for study specific blood work up and biobanking. Data on routine blood and CSF investigations performed as part of the diagnostic work up were collected from the National Health Laboratory Database and patient medical record. The outcome of admission (including death and final diagnosis) were recorded retrospectively.

### Laboratory methods to detect SARS-CoV2 in CSF

An in-house RT-PCR that compared well with the routine nationally employed test, the Multiplex TaqMan™2019-nCoV kit (Applied Biosystems, Waltham, Massachusetts, USA) was established and used to detect SARS-CoV2 in CSF targeting the E gene and subgenomic RNA (sgRNA). Viral subgenomic mRNA is transcribed only in infected cells and not packaged into virions and therefore a positive sample may indicate evidence of actively infected cells within the CSF[16]. RNA was extracted from patient samples using the E.Z.N.A. Viral RNA kit (Omega Bio-tek), followed by reverse transcription and PCR-amplification of the SARS-CoV2-specific targets E and sgRNA (as well as a human RNA control, RNAseP (RP)) using the TaqPath™ 1-Step Master Mix kit (Thermo Fisher) on a QuantStudio 7 Real-Time PCR machine (Thermo Fisher). Primers and probes for SARS-CoV2 E gene and sgRNA readouts have been published elsewhere [17] and were synthesized by Inqaba Biotec (South Africa), while primers and probes for the RP control target were provided by the CDC 2019-Novel Coronavirus (2019-nCoV) Real-Time RT-PCR Diagnostic Panel. Each run included a positive control (PC), which for sgRNA runs included a sample from a previously positive patient, a human specimen extraction control (HSC; e.g. HeLa cell RNA) as well as a no template control (NTC, i.e. water). All samples were amplified under the same conditions using 400nM concentrations of each of the primers, as well as 200nM of probe. Thermal cycling involved 10min at 53°C for reverse transcription, followed by 3min at 95°C to deactivate Reverse Transcriptase and Taq activation, and 45 cycles of 3s at 95°C and 30s at 57°C. A run was considered valid if the control samples yielded the following results with a Ct value < 40 being considered a positive signal: NTC negative for E, sgRNA and RP; HSC negative for E and sgRNA, positive for RP; PC: positive for E, sgRNA and RP. When all controls exhibited the expected performance, an unknown patient sample was considered negative if the Ct values for E and sgRNA were > 37 AND the Ct value for RP < 37. A specimen was considered positive if the Ct values for E and sgRNA were < 37. Runs with a CT between 37 and 40 were repeated.

### Statistical analysis

Data was analysed as an entire cohort with continuous characteristics described in terms of median values and interquartile ranges, and dichotomous variables as counts and percentages. Comparison between patients who tested positive for SARS-CoV2 on nasopharyngeal (NP) swab PCR, and those who did not was assessed using Wilcoxon rank-sum tests to compare continuous variables, and chi squared test for dichotomous variables. All analysis was performed within GraphPad Prism (version 9, Prism for MacOS) software.

## Results

At total of 40 participants (24 female, 16 male) were screened for inclusion in the study. One participant was not enrolled as neither deferred and proxy consent were available. Therefore, 39 participants were included within the analysis. The median age at enrollment was 44 years. Baseline characteristics are outlined in table 1. At the time, routine testing for SARS-CoV2 by NP swab was not available for all inpatients due to limited resource; however, 31/39 participants included within this study underwent testing due to symptomatic presentation, or recent COVID-19 contact. 7/31 participants tested positive for SARS-CoV2 via NP swab. In tables 1 and 2, demographics and clinical characteristics of the cohort are described, as well as for those who tested positive and negative for SARS-CoV2 via NP swab.

**Table 1.** Baseline characteristics.

|  | All | SARS-CoV2 -ve on NP swab or unknown SARS-CoV2 status | SARS-CoV2 +ve on NP swab |
| --- | --- | --- | --- |
| n | 39 | 32 | 7 |
| Gender |  |  |  |
| Male | 15 | 11 | 4 |
| Female | 24 | 21 | 3 |
| Age (years) |  |  |  |
| Median (range) | 44 (27-84) | 42 (27-79) | 47 (42-84) |
| HIV seropositivity |  |  |  |
| HIV co-infected | 22 | 19 | 3 |
| On ART | 11 | 8 | 3 |
| ART naïve/defaulted | 11 | 11 | 0 |
| HIV uninfected | 13 | 11 | 2 |
| Not known | 4 | 2 | 2 |
| HIV-1 VL (copies/ml) (n=13, median ; IQR) | 75831; 28,0141 | 253,344; 258,249 | 205; 206 |
| CD4 Cell count (cells/mm3) (nr 332-1642) (n=16, median; IQR) | 35.5; 186 | 32; 99 | 260; 69 |
| Comorbidities |  |  |  |
| Cardiovascular Disease | 2 | 2 | 0 |
| Hypertension | 13 | 9 | 4 |
| Diabetes Mellitus | 4 | 4 | 2 |
| Obesity | 15 | 11 | 4 |
| Underlying Respiratory Disease | 5 | 5 | 0 |
| Chronic Kidney Disease | 1 | 0 | 1 |
| Baseline Symptoms |  |  |  |
| Cough | 17 | 12 | 5 |
| Fever | 7 | 3 | 4 |
| Dyspnoea | 11 | 7 | 4 |
| Lethargy | 10 | 9 | 1 |
| Nausea | 3 | 3 | 0 |
| Diarrhoea | 11 | 10 | 1 |
| Headache | 12 | 11 | 1 |
| Neck Stiffness | 14 | 13 | 1 |
| Seizures | 7 | 6 | 1 |
| Agitation/psychosis | 10 | 9 | 1 |
| Confusion | 27 | 23 | 4 |
| Anosmia/Dysgeusia | 1 | 0 | 1 |
| Days symptomatic median (IQR) | 7 (13) | 8 (13) | 7 (5) |
| Baseline Neurological Examination |  |  |  |
| Hemiplegia | 5 | 4 | 1 |
| Paraplegia | 1 | 0 | 1 |
| Cerebellar ataxia | 0 | 0 | 0 |
| Tremor | 0 | 0 | 0 |
| Seizures | 1 | 1 | 0 |
| Cranial Nerve Abnormality | 4 | 4 | 0 |
| Glasgow Coma Score (Median) | 14 | 14 | 14 |

**Table 2:** Blood and CSF analysis.

|  | Value expressed as | Performed in (n) | Normal range | All participants | SARS-CoV2 -ve on NP swab or unknown SARS-CoV2 status | SARS-CoV2 +ve on NP swab | p values |
| --- | --- | --- | --- | --- | --- | --- | --- |
| Blood |  |  |  |  |  |  |  |
| Hemoglobin (g/dL) | Median; IQR | 39 | 12-15 | 11.5; 4 | 11.1; 2 | 12.3; 2 |  |
| Platelets (cells x10<sup>9</sup>/L) |  | 37 | 186-454 | 281; 140 | 281; 116 | 271; 168 |  |
| White cells (cells x10<sup>9</sup>/L) |  | 39 | 3.9 – 12.6 | 8.4; 9 | 8.3; 7 | 8.4; 9 |  |
| Neutrophils (cells x10<sup>9</sup>/L) |  | 15 | 1.6 – 8.3 | 6.0; 0 | 7.8; 7 | 4.4; 1 |  |
| Lymphocytes (cells x10<sup>9</sup>/L) |  | 15 | 1.4 – 4.5 | 1.12; 12 | (In 13/15), 1.12; 1 | (in 2/15), 8.56; 8 |  |
| Eosinophils (cells x10<sup>9</sup>/L) |  | 15 | 0.0 – 0.4 | 0.02; 0 | 0.02; 0 | 0.01; 0 |  |
| C-Reactive Protein (mg/L) |  |  | <10 | 68; n/a | 47; n/a | 110; n/a | p=0.36 |
| Erythrocyte Sedimentation Rate (mm/hr) |  | 5 | 0-10 | 96; n/a | 96; 67 | n/a; n/a |  |
| Ferritin (µg/L) |  | 3 | 11-307 | 83; n/a | 83; 492 | n/a; n/a |  |
| D Dimer (ng/mL) |  | 14 | 0 – 0.25 | 1.25; 2 | 1.32; 1 | 0.66; 2 |  |
| ALT (IU/l) |  | 32 | 19-25 | 29.5; 15 | 32; 27 | 19; 16 |  |
| Total Bili (µmol/L) |  | 32 | 5-21 | 12; 8 | 11; 12 | 15; 6 |  |
| Na+ (mmol/L) |  | 39 | 136-145 | 136; 9 | 135; 10 | 137; 6 |  |
| K+ (mmol/L) |  | 39 | 3.5-5.1 | 3.9; 0 | 4.0; 1 | 3.8; 0 |  |
| Creatinine (µmol/L) |  | 39 | 49-90 | 79.0; 24 | 82.5; 64 | 72.0; 18 |  |
| Total Protein (g/L) |  | 24 | 6-78 | 75.0; 14 | 75.0; 13 | 76.5; 10 |  |
| HbA1C (%) |  | 7 | 4-5.6 | 7.9; 6 | 7.25; 2 | 13.0; 0 | p=0.02 |
| Cerebrospinal Fluid |  |  |  |  |  |  |  |
| Polymorphonuclear cells (cells/µL) | Median; IQR; range | 38 | <3 | 0; 6; 0-118 | 0; 1; 0-118 | 0; 0; 0-0 | p=0.39 |
| Lymphocytes (cells/µL) |  | 38 | <3 | 0; 6; 0-2073 | 0; 9; 0-2073 | 2; 4; 0-12 | p=0.62 |
| Erythrocytes (cells/µL) |  | 38 | <3 | 5; 48; 3-349 | 3; 17; 0-349 | 62; 148; 0-311 | P=0.28 |
| Protein (g/L) |  | 39 | 0.15-0.45 | 0.39; 1; 0.13-4.82 | 0; 1; 0.13 – 4.82 | 0; 0; 0.17 – 2.19 | p=0.56 |
| Glucose (mmol/L) |  | 39 |  | 3; 3; 0.1-10.2 | 3; 2; 0.1-7.0 | 5; 5; 2.8 – 10.2 | p=0.01 |
| Bacterial Culture | n, (% positive) | 39 |  | 5/39 (12.8) | 4/32 (12.5) | 1/7 (14.2) | p=0.90 |
| CLAT | n, (% positive) | 32 |  | 5/32 (15.6) | 5/29 (17.2) | 0/3 (0) |  |
| GXPU | n, (% positive) | 22 |  | 1/22 (45.5) | 1/18 (5.5) | 0/4 (0) |  |
| PCR for SARS-CoV2 in CSF |  |  |  |  |  |  |  |
| Positive CT value for E |  | 39 |  | 0/39 | 0/32 | 0/7 |  |
| Positive CT value for sgRNA |  | 39 |  | 0/39 | 0/32 | 0/7 |  |

Values expressed as Median;Interquartile Range (IQR); Range, or n (% positive). Abbreviations: Na+: sodium; K+: potassium; CLAT: cryptococcal latex agglutination titre; GXPU: GeneXpert Ultra; CT (cycle threshold); sgRNA: subgenomic RN.

Neurological complaints at baseline are described in table 1. The most complaint was confusion (27/39). Less common complaints, occurring with or without confusion included: neck stiffness (14/39), headache (12/39), new onset or increasing frequency of seizures (7/39), acute psychotic symptoms (10/39). On neurological assessment, 5/39 had new onset hemiplegia and 1/39 had new onset bilateral lower limb weakness. In the absence of other motor or sensory disturbance, 1/39 had new onset lower motor neuron facial nerve (VII cranial nerve) weakness, and 1/39 had abducens (VI cranial nerve). Neurological complaints at presentation in those testing positive for SARS-CoV2 on NP swab included: confusion (4/7) of which 1/7 demonstrated acute psychotic symptoms, new onset seizures (1/7), headache and neck stiffness (1/7), acute onset right sided weakness (1/7) and bilateral lower limb weakness (1/7). Of these 7 participants, 3 had no classical symptoms of COVID-19 (cough, shortness of breath, fever, anosmia or dysgeusia). Between those with and without a positive NP swab for SARS-CoV2, there was no significant difference in the neurological complaints at baseline. In 7 patients with a confirmed diagnosis of COVID-19 (SARS-CoV2 positive PCR on NP swab); 2/7 were thought to have COVID-19 pneumonia with stroke (see table 4), 1/7 presented with a clinical diagnosis of myelitis treated as possible viral or *M. tb* in aetiology, with the remaining 4/7 patients, all of which presented with confusion, thought to be due to delirium secondary to COVID-19 pneumonia. No further investigation was performed to formally assess for encephalopathy.

Baseline blood and CSF analysis are described in table 2. This analysis was performed as part of routine care, where there was a clinical indication and therefore not all tests were performed on every participant. There was no difference in CSF markers to suggest an acute infective or inflammatory process in those who tested positive for SARS-CoV2 via NP swab, versus those who did not, this includes: lymphocyte count (*p=0*.*62*), polymorphonuclear cells (*p=0*.*39*) and protein (*p=0*.*59*). CSF glucose was significantly higher in those with a diagnosis of COVID-19 (5.0 vs 3.0, *p=0*.*01*); however, of note HbA1C (%) was significantly higher in patients with COVID-19 than in those without (13.0 vs 7.25, *p=0*.*02*), reflecting the non-significant higher proportion of patients with pre-existing diabetes mellitus in those with a diagnosis of COVID-19 compared to those without (2/7 vs 4/32, *p=0*.*29*).

Computerised Tomography (CT) scans of the brain were performed at baseline in 26/39 participants, of which 2 were performed in patients with a confirmed diagnosis of COVID-19. 3/26 scans were performed with contrast enhancement. Radiological findings at baseline are summarized in table 3. In the two scans performed in patients with COVID-19, one demonstrated multi-focal subacute infarcts within the left middle cerebral artery (MCA) and anterior deep borderzone territories. In the second, imaging demonstrated multi-territory mature infarcts in in both cerebral hemispheres and in the cerebellum (figure 1). In both patients, CSF findings were unremarkable (see table 4), and a diagnosis of COVID-19 pneumonia with presumed diagnosis of stroke (clinical in the former, radiological in the latter) was made.

**Table 3:** Radiological analysis.

| Radiological Finding | All participants | SARS-CoV2 -ve on NP swab or unknown SARS-CoV2 status | SARS-CoV2 +ve on NP swab |
| --- | --- | --- | --- |
| Leptomeningeal Enhancement | 1/26 | 1/24 | 0/2 |
| Hydrocephalus | 3/26 | 3/26 | 0/2 |
| of which communicating | 3/26 | 3/26 | n/a |
| Radiological evidence of infarcts | 14/26 | 13/26 | 1/2 |
| of which new | 3/14 | 3/13 | 0/1 |
| single | 10/14 | 9/13 | 1/1 |
| multiple | 4/14 | 4/13 | 0/1 |
| Territory |  |  |  |
| Middle cerebral artery territory | 9/14 | 8/13 | 1/1 |
| Anterior cerebral artery territory | 0/14 | 0/13 | 0/1 |
| Posterior cerebral artery territory | 0/14 | 0/13 | 0/1 |
| Ring enhancing lesions | 0/26 | 0/24 | 0/2 |
| Effacement | 1/26 | 1/24 | 0/2 |
| Of which local sulcal | 1/26 | 1/24 | 0/2 |
| Of which hemispheric sulcal | 1/26 | 1/24 | 0/2 |
| Of which global sulcal | 0/26 | 0/24 | 0/2 |
| Of which basal cistern | 0/26 | 0/24 | 0/2 |

**Table 4:**
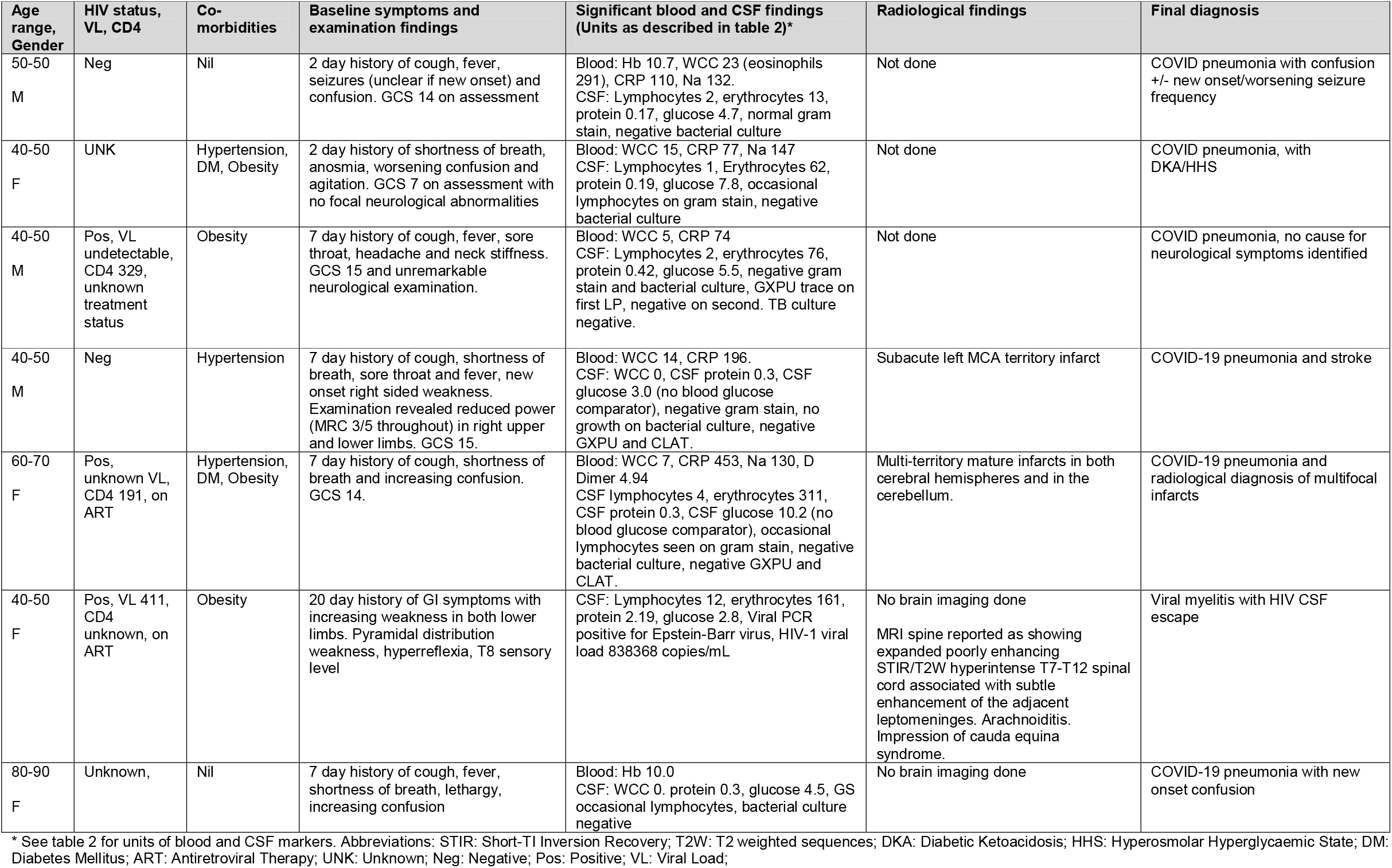
Detailed clinical presentations, laboratory findings in patients presenting with COVID-19 and neurological symptoms.

| Age range, Gender | HIV status, VL, CD4 | Co-morbidities | Baseline symptoms and examination findings | Significant blood and CSF findings (Units as described in table 2)* | Radiological findings | Final diagnosis |
| --- | --- | --- | --- | --- | --- | --- |
| 50-50<br>M | Neg | Nil | 2 day history of cough, fever, seizures (unclear if new onset) and confusion. GCS 14 on assessment | Blood: Hb 10.7, WCC 23 (eosinophils 291), CRP 110, Na 132.<br>CSF: Lymphocytes 2, erythrocytes 13, protein 0.17, glucose 4.7, normal gram stain, negative bacterial culture | Not done | COVID pneumonia with confusion +/- new onset/worsening seizure frequency |
| 40-50<br>F | UNK | Hypertension, DM, Obesity | 2 day history of shortness of breath, anosmia, worsening confusion and agitation. GCS 7 on assessment with no focal neurological abnormalities | Blood: WCC 15, CRP 77, Na 147<br>CSF: Lymphocytes 1, Erythrocytes 62, protein 0.19, glucose 7.8, occasional lymphocytes on gram stain, negative bacterial culture | Not done | COVID pneumonia, with DKA/HHS |
| 40-50<br>M | Pos, VL undetectable, CD4 329, unknown treatment status | Obesity | 7 day history of cough, fever, sore throat, headache and neck stiffness. GCS 15 and unremarkable neurological examination. | Blood: WCC 5, CRP 74<br>CSF: Lymphocytes 2, erythrocytes 76, protein 0.42, glucose 5.5, negative gram stain and bacterial culture, GXPU trace on first LP, negative on second. TB culture negative. | Not done | COVID pneumonia, no cause for neurological symptoms identified |
| 40-50<br>M | Neg | Hypertension | 7 day history of cough, shortness of breath, sore throat and fever, new onset right sided weakness. Examination revealed reduced power (MRC 3/5 throughout) in right upper and lower limbs. GCS 15. | Blood: WCC 14, CRP 196.<br>CSF: WCC 0, CSF protein 0.3, CSF glucose 3.0 (no blood glucose comparator), negative gram stain, no growth on bacterial culture, negative GXPU and CLAT. | Subacute left MCA territory infarct | COVID-19 pneumonia and stroke |
| 60-70<br>F | Pos, unknown VL, CD4 191, on ART | Hypertension, DM, Obesity | 7 day history of cough, shortness of breath and increasing confusion. GCS 14. | Blood: WCC 7, CRP 453, Na 130, D Dimer 4.94<br>CSF lymphocytes 4, erythrocytes 311, CSF protein 0.3, CSF glucose 10.2 (no blood glucose comparator), occasional lymphocytes seen on gram stain, negative bacterial culture, negative GXPU and CLAT. | Multi-territory mature infarcts in both cerebral hemispheres and in the cerebellum. | COVID-19 pneumonia and radiological diagnosis of multifocal infarcts |
| 40-50<br>F | Pos, VL 411, CD4 unknown, on ART | Obesity | 20 day history of GI symptoms with increasing weakness in both lower limbs. Pyramidal distribution weakness, hyperreflexia, T8 sensory level | CSF: Lymphocytes 12, erythrocytes 161, protein 2.19, glucose 2.8, Viral PCR positive for Epstein-Barr virus, HIV-1 viral load 838368 copies/mL | No brain imaging done<br>MRI spine reported as showing expanded poorly enhancing STIR/T2W hyperintense T7-T12 spinal cord associated with subtle enhancement of the adjacent leptomeninges. Arachnoiditis. Impression of cauda equina syndrome. | Viral myelitis with HIV CSF escape |
| 80-90<br>F | Unknown, | Nil | 7 day history of cough, fever, shortness of breath, lethargy, increasing confusion | Blood: Hb 10.0<br>CSF: WCC 0. protein 0.3, glucose 4.5, GS occasional lymphocytes, bacterial culture negative | No brain imaging done | COVID-19 pneumonia with new onset confusion |

\* See table 2 for units of blood and CSF markers. Abbreviations: STIR: Short-TI Inversion Recovery; T2W: T2 weighted sequences; DKA: Diabetic Ketoacidosis; HHS: Hyperosmolar Hyperglycaemic State; DM: Diabetes Mellitus; ART: Antiretroviral Therapy; UNK: Unknown; Neg: Negative; Pos: Positive; VL: Viral Load;

**Figure 1:**
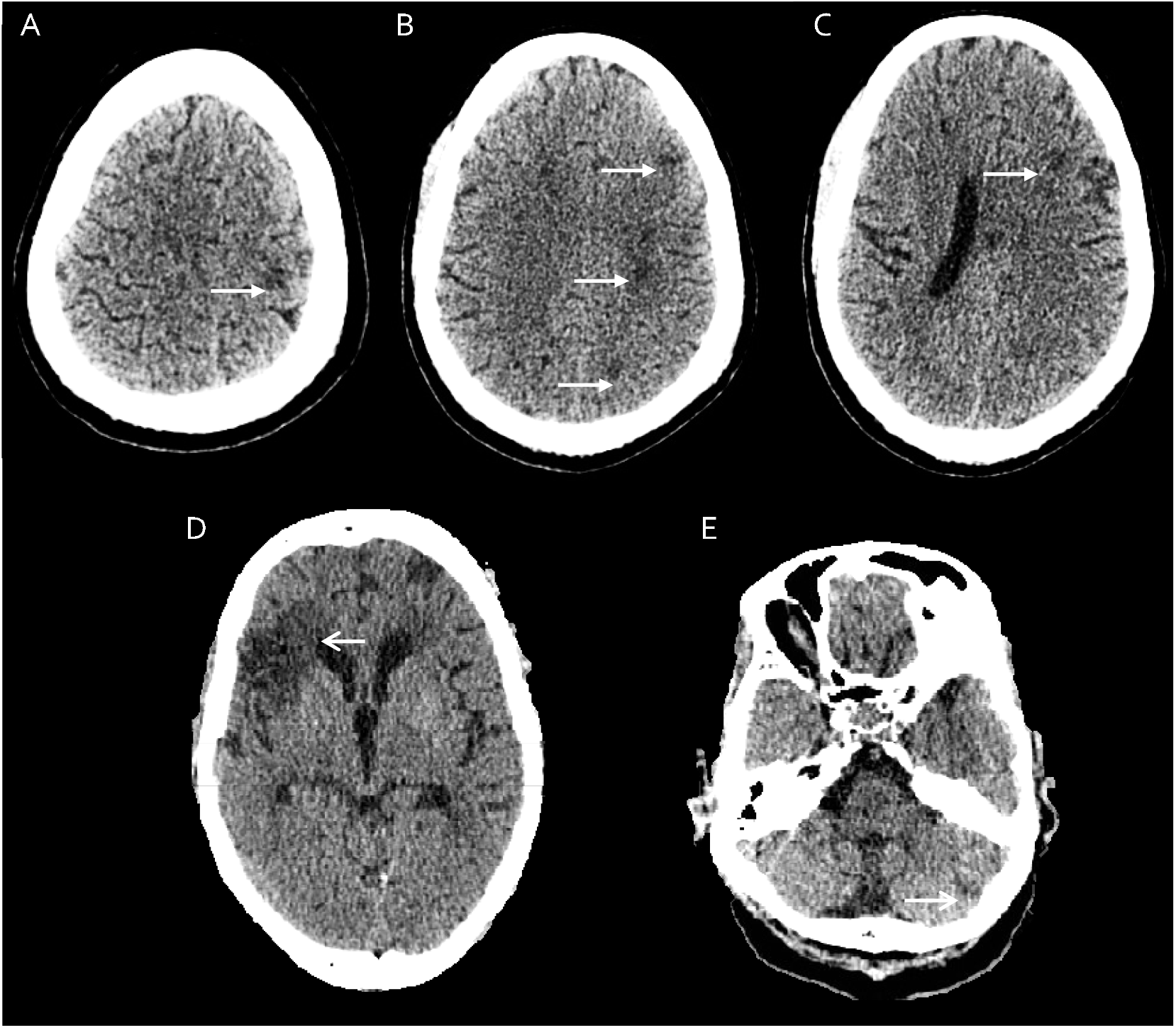
Axial unenhanced CT head imaging in two patients where a diagnosis of COVID-19 was confirmed (PCR for SARS-CoV2 positive on NP swab). Top row demonstrates poorly-defined multifocal cortical and subcortical hypodensities in keeping with subacute left middle cerebral artery territory (A,C) a anterior deep borderzone territory (B) infarcts. Bottom row demonstrates multi-territory mature infarcts in the right middle cerebral artery terr tory (D) and l posterior inferior cerebellar artery territory (E).

Multiplex PCR (targeting nucleocapsid (N), spike protein (S) and Orf1ab) runs revealed no evidence of SARS-CoV2 in any of the 39 samples. Using the primer combinations for E gene readout and subgenomic readout repeatedly gave no or weak (Ct>37) signals that were interpreted as negative. Therefore, none of the 39 samples demonstrated SARS-CoV2 via any of the PCR primer combinations applied. Raw data is presented in table 1 of the supplementary material.

## Discussion

We describe a cohort of patients presenting with clinical symptoms suggestive of possible neuroinfective or neuroinflammatory aetiology, with and without symptoms and a confirmatory diagnosis of COVID-19 during the first peak of the pandemic in a resource poor setting in South Africa. Examination of the CSF using PCR for multiple targets was negative in all cases therefore suggesting little evidence of direct neurotropic invasion of SARS-CoV2 in CNS.

Published reports provide rationale for direct neurotropic invasion of SARS-CoV2 including cases of meningitis and encephalitis where PCR for SARS-CoV2 was positive in CSF with and without classical symptoms of COVID-19 [2] [[18]. At autopsy, SARS-CoV2 RNA has been detected in the brains of patients who have died due to COVID-19 albeit at titers lower than in other affected organs [19, 20]. Evidence for direct neurotropic invasion is supported by findings from the 2002 SARS-CoV outbreaks where studies demonstrated the presence of coronavirus particles in the brain [21-23], with subsequent studies describing penetration of the CNS via the olfactory nerve [24]. In SARS-CoV2, a case of olfactory gyrus intracerebral hemorrhage, an uncommon location for spontaneous hemorrhage, as well as the high rates of anosmia, has highlighted whether SARS-CoV2 can invade neurological structures such as the olfactory bulb via nasal mucosa [25]. Entry into the CNS via synaptic connections may also provide rationale to consider a centrally-driven contribution to cardiorespiratory dysfunction in coronaviruses, where acute onset respiratory failure leads to significant morbidity and mortality [36]. These observations have, during the course of the COVID-19 pandemic, raised questions as to whether SARS-CoV2 should be investigated as a causative organism, particularly in patients presenting with a meningitis or encephalitis in a setting where SARS-CoV2 infection rates are high, both in those with and without a confirmed diagnosis of COVID-19. In our cohort who were enrolled sequentially in a tertiary setting for COVID-19 care, despite detailed examination of the CSF for presence of SARS-CoV2 using PCR primers for genomic and subgenomic RNA, no patients were found to have evidence of direct neurotropic invasion to the CNS. This finding is important in shaping the direction of clinical care in the management of patients presenting with neurological symptoms in the COVID-19 era, particularly in resource limited settings where the prioritisation of investigations is an important consideration in clinical care.

In our study, one patient had a clinical presentation consistent with myelitis alongside a diagnosis of COVID-19, however in this case an explanation other than SARS-CoV2 infection was thought more likely to account for their clinical presentation, and no evidence of direct neurotropic invasion of SARS-CoV2 was found. Acute myelitis [26] is one of many cases reported in the literature where the mechanism was thought due to a post or para-infectious inflammatory response to SARS-CoV2. Other cases which may suggest inflammatory sequelae occurring during or following SARS-CoV2 infection include: acute necrotising hemorrhagic encephalopathy [27, 28], Guillain Barre Syndrome [10-12, 29] and Miller Fisher Syndrome [30], and acute disseminated encephalomyelitis [31, 32], all of which occurred without evidence of SARS-CoV2 in the CSF. Given that within our cohort direct neurotropic invasion of the CNS was not found, these results might suggest that greater emphasis should now turn towards understanding the role of inflammation at the time of or following SARS-CoV2 infection in a subset of patients.

Moreover, the proportional contribution of the now well-described coagulopathy leading to endothelial dysfunction and eventual end organ damage is unknown [33]. This is particularly important to understand in the context of stroke in patients with COVID-19; now frequently reported to occur where otherwise no clear vascular risk factors exist [34, 35]. In our cohort, two patients presented with presumed stroke alongside COVID-19 pneumonia. In both cases, vascular risk factors co-existed and may in part or completely explain the vascular complications. Further research is required to understand the interplay of the presumed coagulopathy both on pre-existing vascular risk factors such as hypertension and diabetes, and other stroke risk factors such as HIV, particularly within the South African context.

There were several limitations to this study. Given the pragmatic nature of its design, only data on investigations indicated as part of routine clinical care were available for analysis, resulting in an incomplete laboratory and radiological data set. This includes cerebral imaging, which was not performed in all participants, and in instances where it was, contrast was not given in the majority (23/26) of cases. Factors related to coagulation, such as D-Dimer levels, would have provided interesting comparison of patients presenting with complications such as stroke with and without confirmed COVID-19. In contrast, laboratory procedures related to the discovery of SARS-CoV2 in CSF were thorough and robust, which reassures that despite multiple runs, the negative findings are reliable.

Although small, this pragmatic observational cohort study contributes knowledge to our increasing understanding of COVID-19 management. Through systematic analysis of CSF in patients presenting with neurological symptoms in a context where incidence of SARS-CoV2 infection is high we have demonstrated that although cases within the literature exist, direct neurotropic invasion of the CNS is uncommon. This includes suspected cases of meningitis and encephalitis, syndromes most aligned to direct neurotropic mechanism. This considered, neurological presentations in cases of COVID-19 continue to be reported, and lead to morbidity and mortality in patients affected. The results from our study suggest that the further emphasis must now turn towards understanding the role of inflammation and coagulopathy in the development of neurological syndromes. This includes studies to assess the efficacy of proven anti-inflammatory drugs such as corticosteroids and tocilizumab, and therapeutics to manage acute stroke in the treatment of patients who develop neurological symptoms due to SARS-CoV2 infection.

## Data Availability

Data archived

## Supplementary Material

**Table 1:**
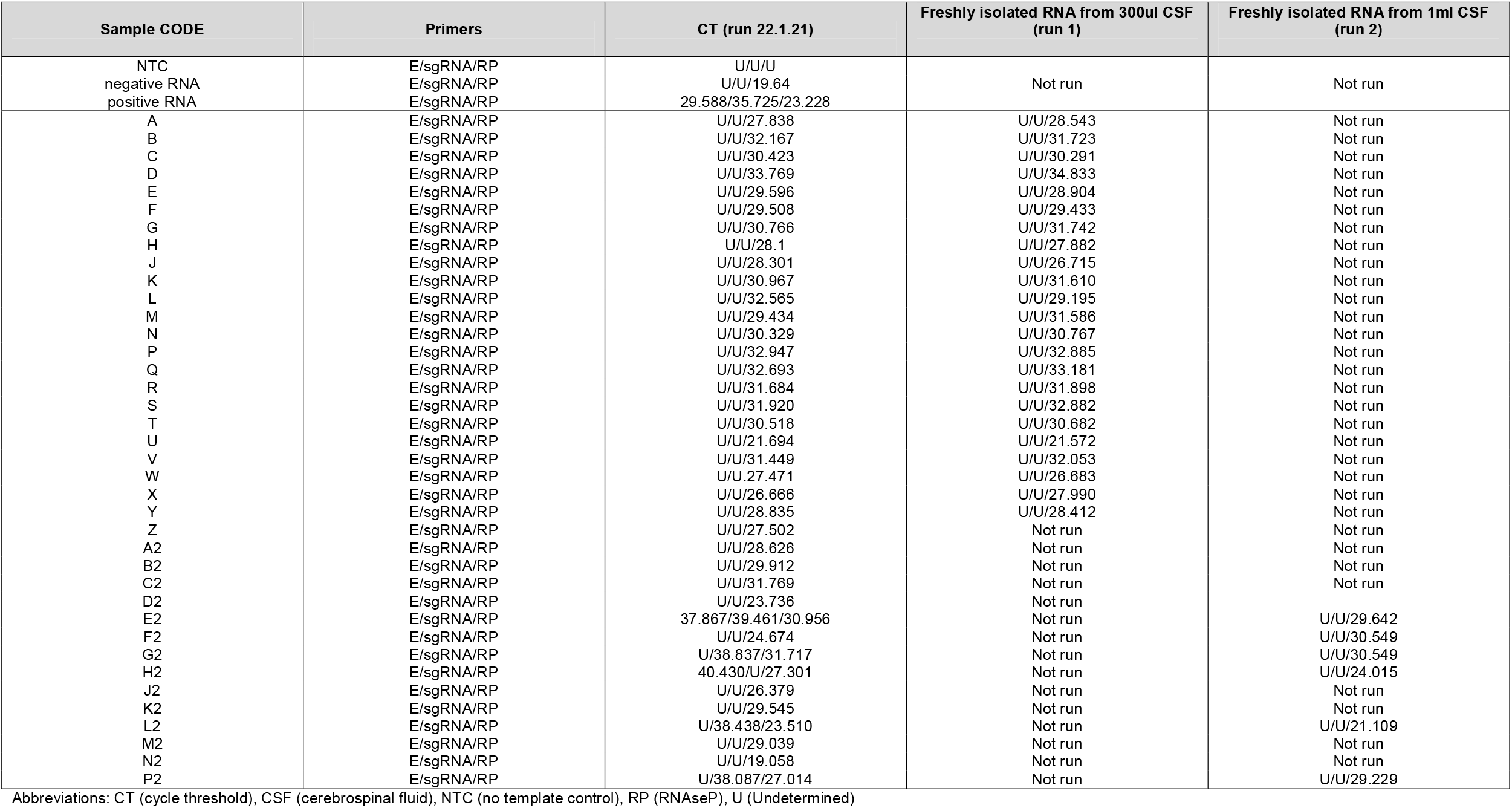
Results of CSF PCR analysis for detection of SARS-CoV2 (Supplementary data)

